# Staff perceptions of an NHS military veterans’ mental health service in the UK: A qualitative evaluation

**DOI:** 10.64898/2026.10.06.26364875

**Authors:** Shivangi Talwar, Carlos Osório, Emma Barslund Blackman, Sacha Solomons, Jaahnvi Krishnamurthy, Ronja Kuhn, Leon Culloty, Talya Greene, Sue Ferrier, Jo Billings

## Abstract

Those leaving military service frequently present with complex psychosocial problems, but military veterans are often reluctant to engage with statutory healthcare provision. Specialist mental health services have recently been established in the UK to specifically support military veterans with their psychological and social wellbeing. We explored the treatment and support provided by the London branch of a military veterans’ mental health and wellbeing service (Op COURAGE) from the perspectives of the multidisciplinary service team and their partner organisations and collaborators. We conducted semi-structured interviews with multidisciplinary service staff and members of their partner organisations and external agencies. Qualitative data were analysed using framework analysis. A total of 16 professionals participated in this study, comprising five service staff, eight collaborators and three partner agency participants. Five themes with subthemes were identified, including (1) blended networks, (2) staff inclusivity and sensitivity, (3) spectrum of psychosocial interventions and holistic support, (4) referral mechanisms, and (5) (in)visibility of the service and advertising. Our evaluation highlighted the advantages of a multidisciplinary team and collaborative partnerships to meet the psychosocial needs of veteran and their significant others. We have proposed that service improvement could include increasing advertisement about this specialist service, deepening knowledge about veterans among staff and further involving veterans and key stakeholders in service decisions.

## Introduction

Military veterans have been consistently reported to experience higher rates of mental health problems than the general civilian population [1, 2]. In the UK, a recent longitudinal study found that the prevalence of common mental health disorders (CMDs) amongst military veterans was 27.9%, probable Post-Traumatic Stress Disorder (PTSD) 10.5%, Complex PTSD 6.5%, and alcohol misuse 8.7% [3]. While these findings seem to indicate relatively high levels of mental health problems, research also shows that most veterans do not seek support for their difficulties [4]. Reported barriers in engaging UK veterans include stigma, lack of awareness, lack of veteran-specific services and gender disparities [5, 6]. Whilst veterans may have awareness of physical health support structures, they may still lack knowledge of the need for, or availability of, mental health services. The House of Commons Defence Committee’s (2019) efforts to improve mental healthcare for veterans in the UK led to a tri-model service and the development of Op COURAGE mental health and well-being services across the UK since April 2023 [7].

The Op COURAGE service model includes a multidisciplinary team offering interventions using psychological, social, pharmacological and nursing approaches as well as veteran peer support [8]. In England, there are a total of six Op COURAGE services [9]. This paper reports on the study conducted as part of an independent evaluation of the London branch of the Op COURAGE service, commissioned by NHS England.

The London Op COURAGE service is a collaboration of NHS and third sector providers. The North London NHS Foundation Trust and Oxleas NHS Foundation Trust provide clinical interventions. Within this collaborative, three charity partners (Walking with the Wounded, The Stoll Foundation, and The Ripple Pond) support veterans with social vulnerabilities due to social isolation, benefits and employment. Walking with the Wounded have particular expertise in employment [10]. The Stoll Foundation offers housing support to vulnerable and disabled veterans, including affordable accommodation [11]. The Ripple Pond focuses on the significant others of physically and psychologically wounded veterans by organising support groups and talks and offering help with employment and referrals [12].

We aimed to explore the treatment and support offered at London Op COURAGE from the perspectives of the multidisciplinary service team, partner agencies (i.e., Walking with the Wounded, The Stoll, and Ripple Pond), and external collaborators (i.e. staff at NHS Trusts and third-sector organisations). Through this multi-stakeholder approach, we identified strengths and areas for potential improvement to inform future service development.

## Materials and Methods

### Approvals

We received permission from North London NHS Trust for a service evaluation of the London Op COURAGE Veterans’ Mental Health and Wellbeing service. This project was classed as a Service Evaluation; therefore, no separate NHS ethics application was necessary. Written informed consent was obtained from all participants to take part in the interviews reported in this paper. A research team at University College London (UCL) were commissioned by NHS England to conduct this study as part of an independent evaluation of the London Op Courage service. Throughout the reporting of this qualitative interview study we adhere to the Standards for Reporting Qualitative Research checklist [13].

### Eligibility criteria

We included three groups of professionals working with veterans and their families; multidisciplinary service staff from Op COURAGE, partner agencies’ staff, and external collaborators. The inclusion criteria were as follows:

#### Multidisciplinary staff

- Mental health professionals, support workers and social workers working at the London Op COURAGE service
- Delivering treatments and/or support to the service users within the Op COURAGE service

#### Partner agencies’ staff

- Working at Walking with the Wounded, The Stoll or The Ripple Pond
- Management staff overseeing the support workers at the London Op COURAGE service

#### External collaborators

- Working in NHS Trusts or charity organisations who may refer to Op COURAGE and providing support to users of the Op COURAGE service

### Recruitment of participants

We recruited all participants through clinical staff (mainly SF and RK) at the London Op COURAGE service, between December 2024 and March 2025. Potential participants were invited to take part through email, in person or via study recruitment flyers. ST coordinated the recruitment process with RK, EBB, SS and JK. We sent the participant information sheet and consent form to those who expressed interest. On receiving written informed consent, the assigned interviewer (ST, RK, EBB, SS or JK) set up an interview at a mutually agreed time and location.

### Data collection

ST, RK, EBB, SS and JK conducted semi-structured interviews online via MS Teams or in person at the Op COURAGE service, as convenient for the participant. ST developed the topic guides for the interviews, which were reviewed by JB, SF, DM, LC and CO (also a military veteran) (S1 Topic guide) These questions were also reviewed by UK veteran lived experience advisors. Interview questions explored the experiences of service staff in delivering treatment and care with London Op COURAGE. Partner agencies and external collaborators were asked about their views on the London Op COURAGE staff and facilities, and suggestions for improving the service.

### Data analysis

We used framework analysis to interpret the data (Gale et al., 2013). EBB, SS and JK coded two transcripts from each group of participants to develop the initial framework. ST reviewed the framework and further refined it with EBB, SS and JK. Any relevant data not fitting in the framework was included under ‘other’. The remaining interviews were coded by EBB, SS, JK, ST and RK, who continued to refine the framework with JB. The team met regularly during coding and framework development. ST developed the preliminary themes and subthemes, which were discussed with EBB, SS and JK and refined with JB. All team members reviewed the analysis presented in this manuscript.

### Reflexivity

The research team comprised academics and clinicians at various career stages, with backgrounds in mental health and experience of working with military veterans and with people with PTSD. ST, CO, LC, DM, SF and JB are qualified mental health clinicians. EBB, SS and JK have completed postgraduate-level mental health studies, and RK has research experience with people with mental health conditions. ST, CO, TG and JB specialise in research with people affected by trauma. CO, LC, DM, SF and JB have clinical and research experience specifically with veterans. CO is a former Army Staff Sergeant and has been deployed overseas. ST and JB are experienced qualitative researchers who oversaw the project and supervised EBB, SS, JK and RK. Our varied interests and experiences informed the study design and materials, as well as analysis.

## Results

We interviewed 16 professionals; five service staff, three from partner agencies, and eight external collaborators. Of those, 15 were of White ethnicity and one of Black ethnicity. Ten participants were male, and the remaining six were female. Table 1 describes the characteristics of participants in detail.

**Table 1:** Characteristics of participants.

| Number of Participants | n (16) |
| --- | --- |
| Service/MDT staff | 5 |
| Partner agency staff | 3 |
| External collaborator staff | 8 |
| Mean age (in years) | 51.8 |
| 30-40 | 1 |
| 41-50 | 6 |
| 51-60 | 5 |
| 61-70 | 4 |
| Sex |  |
| Male | 10 |
| Female | 6 |
| Ethnicity |  |
| White British | 13 |
| White Scottish | 1 |
| White Other | 1 |
| Black British | 1 |

The interviews lasted between 15 minutes and 1 hour and were an average of 46 minutes long.

We developed six themes: 1) blended networks, 2) staff inclusivity and sensitivity, 3) holistic and personalised support, 4) approaching the veterans, 5) referral mechanisms and 6) (in)visibility of the service and advertisement. We have conceptualised staff sensitivity, blended networks and holistic and personalised support as ‘within service’ factors and approaching veterans, referral mechanisms and invisibility requiring advertisement as information flows in and out of the service (depicted in Fig 1). Further sub-themes within some of these themes are listed in Table 2 and illustrated below with anonymised quotes.

**Fig 1:**
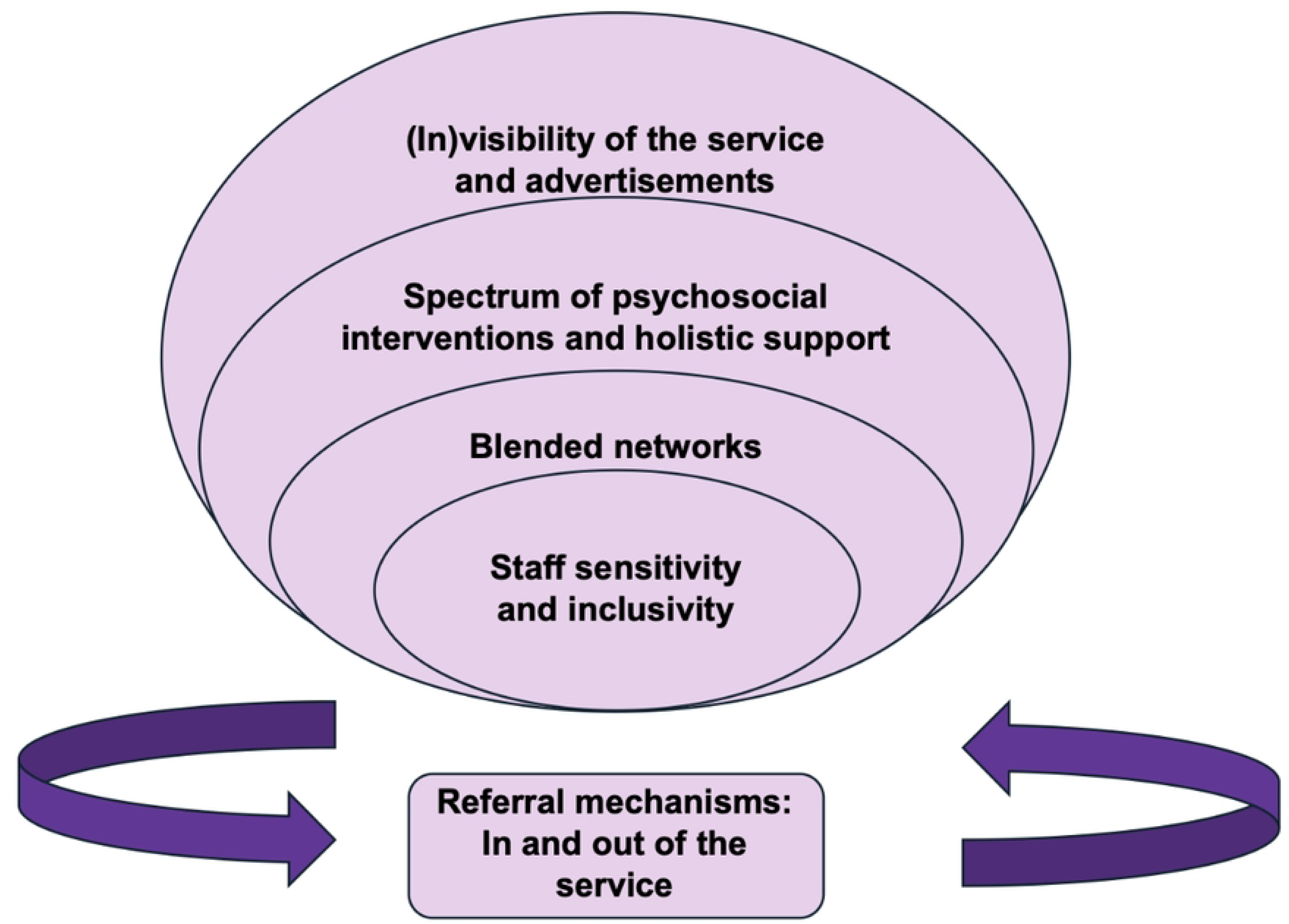
Themes

**Table 2:** Themes and subthemes for service providers’ data.

| Themes | Subthemes |
| --- | --- |
| **1. Blended networks** | 1.1 Developing networks |
|  | 1.2 Highs and lows of interactions with "outsourced" services |
| **2. Staff inclusivity and sensitivity** | 2.1 Evolving veteran sensitivity |
|  | 2.2 Sensitivity towards diverse groups |
| **3. Spectrum of psychosocial interventions and holistic support** |  |
| **4. Referral mechanisms: In and out of the service** | 4.1 (In)convenience of referral forms |
|  | 4.2 Referrals require details |
| **5. (In)visibility of the service and advertisement** |  |

### 1. Blended networks

In this theme, we have captured the within-service interactions and their network of organisations and Trusts.

#### 1.1 Developing networks

All partners and collaborators highlighted their good relations with the service lead and staff members. They emphasised the service staff’s investment in forming a network of organisations, partners and professionals for veterans’ needs. These collaborators appreciated the efforts of the MDT to engage with communities and improve the functioning of other organisations.

> *“they’ve got great links into community, community mental health and so…hopefully no veteran will just be left without treatment.”-External organisation staff*

> *“It’s really good that they’re so open, always have been from the very start, and (an Op COURAGE clinician) has been really instrumental in helping us, you know, develop what we do. So, she’s gone beyond, you know, her remit really.”-External organisation staff*

A partner agency participant expressed the need to be updated on their clients. One external organisation participant also spoke about being unsure about what support is available for the veterans on the service’s waiting list. A few of the partner and collaborator participants emphasised the need for improved networking for veteran care. They suggested that the service expand its collaborations to improve awareness about this specialist service and seamless referrals. The service staff, on the contrary, spoke about their efforts to deliver presentations, engage with diverse groups and enhance their network. They were confident of the connections they had developed with communities and organisations. Service staff also spoke about their extensive efforts to maintain those networks.

> “Gurkha veterans, I do engagement with them, and try and build up relationships and break down stigma”-MDT staff

> *“I also liaise with the different charities across London. And that might include things like a monthly meeting that I will chair with one of the charities, such as xxx Housing-” MDTstaff*

#### 1.2 Highs and lows of interactions with “outsourced” services

London Op COURAGE works collaboratively with other organisations and Trusts within London to meet those veteran needs which are outside of the service’s remit. According to the MDT staff, they make efforts to engage with these partners and collaborators to support their work and offer continued support to the veteran service users. Most partners and external organisation participants echoed this view that they receive mental health consultation and, at times, decision-making and brand value from the service.

> *“we’re on the phone regularly, discussing clients’ behaviours…”-External organisationstaff*

> *“it’s quite top notch in terms of what a collaborative partnership would look like”-External organisation staff*

> *“she’s (a clinician at Op COURAGE) been involved right from the very start and she doesn’t have to…So it’s in addition to what she’s doing as her job. She sat on our steering group. She’s turned up to every meeting..”-External organisation staff*

Some participants from partner agencies attended Op COURAGE’s MDT meetings as well as invited the service staff to theirs. This regular contact ensured that the intent to offer holistic care is communicated to others supporting a particular veteran. However, some of the partners and collaborators suggested regular meetings or annual updates.

> *“it’s always helpful to do a bit of an annual update. Definitely, because I think even low level things change within the service or personnel change or slight structures change or the offer changes.”-External organisation staff*

Most partners and collaborators said that they did not need to know the role of all MDT staff. However, understanding the service structure at regular intervals or annually would be beneficial. The collaborators, specifically, were sometimes unsure of the service structure and staff turnover. One of these partner agency participants reported that they met a new clinician during every interaction, highlighting potential inconsistencies and difficulties in maintaining lasting professional relationships.

> *“I’m always meeting someone new from Op COURAGE. I don’t know if it’s turnover or just lots of different areas I haven’t known, but every time I meet up, there’s another face.”-External organisation staff*

### 2. Staff inclusivity and sensitivity

In this theme, we have described the service staff’s sensitivity towards this veteran population and awareness of military life. We have further elaborated on suggestions to improve this inclusive and sensitive approach.

#### 2.1 Evolving veteran sensitivity

All participants, from all the groups, highlighted the awareness and sensitivity of the London Op COURAGE service staff towards veteran needs. Partners and collaborators cumulatively voiced that the service staff are veteran-sensitive and that their veteran staff members are invaluable. One participant also highlighted that the leadership has been consistently working with this population, ensuring reflective practice and service progression.

> *“the leadership..They’ve certainly got the longevity of working with the service and evolving the service as it’s gone forward, which has been really great and having that continuity of leadership is also really, I think is also really important for the service that you’ve got a couple of people at the top that have been here for ages…actually they then can be reflective on what’s happened in the past……But equally, be progressive about what moves forward…very veteran responsive service”-External organisation staff*

MDT participants emphasised that they are veteran aware and that they make efforts to improve their understanding. They said they make special efforts to ensure that they remain approachable.

> *“I always make it clear to people—it’s always open. They can always come back to the service if they need help. We are an approachable service. They know my phone number, my email. If something comes up on the social side of things, I can give them pointers and advise them on where to go to resolve it.”-MDT staff*

A few of the participants suggested that the lack of military background of some of the staff members could be an obstacle to forming connections with veteran service users. One participant highlighted the need for training staff, especially new members, about veteran life and ranks. Another participant suggested continued training by specialists, including academic teams.

> *“I feel the general service would benefit from, especially with staff turnover, some basic training for clinicians around veteran awareness training…and social welfare issues, just to raise their understanding with the processes a little more. Service users will talk about their problems, and clinicians may not quite understand.”-MDT staff*

#### 2.2 Sensitive towards diverse groups

Most participants highlighted the inherent socio-demographic diversity in London, leading to the diverse composition of service users from different communities. Some of the partners and collaborators suggested that there is a need to visit veterans from different communities to understand their needs and enable them to improve access to professional help. A few partners and collaborators emphasised that some members of the MDT take special measures to engage with groups like LGBTQ and Gurkhas.

> *“there’s very strong links with military charities and Fighting with Pride. Very strong links with the Foreign Commonwealth cultures and with obviously with Gurkha welfare trust, so they’ve got good links across.”-External organisation staff*

### 3. Spectrum of psychosocial interventions and holistic support

This theme describes London Op COURAGE meeting a wide array of veteran needs and developing networks to meet those needs. Our participants described the diverse range of mental health and social interventions available within the service. These included psychological and pharmacological interventions, help with housing and employment, peer support and carers’ consultations. They emphasised the varied treatment approaches offered by the service staff and specialised training in those approaches.

> *“…catalogue of different interventions…a lot of the staff are trained in many different modalities…so that might include things like DBT, CBT, EMDR, CFT, ACT, systemic stuff, systemic therapy, and some psychodynamic stuff…there’s people doing interpersonal therapy, and we’ve been trained in mentalisation-based therapy as well and other things like that”-MDT staff*

Specifically, external collaborators reported that they were unclear about the “different levels of support” and the nuances of the facilities. They, however, said that being less aware of those details did not impact their collaboration. Further, external collaborators and partner agency staff showed confidence in the skills of the MDT. They conveyed the reliability of the service to support the mental health of veterans.

> *“it would be my personal first recommendation for anybody who is a veteran that needs mental health support. I would say try Op COURAGE first.”-External organisation staff*

A few participants highlighted the drop in and outreach work by London Op COURAGE. A collaborator spoke about coordination with a service staff member regarding five homeless veterans within two hours. However, some collaborators and partners suggested that there was scope to enhance outreach work, especially for communities where mental health care is stigmatised.

> *“with Pride and the LGBTQ community, there’s been a lot of positive work done”-MDT staff*

> *“I think there needs to be lots of videos, lots of outreach.”-External organisation staff*

### 4. Referral mechanisms: In and out of the service

In this theme, we have captured the experiences of professionals in using the referral process followed by the service staff.

#### 4.1 (In)convenience of referral forms

Some of the MDT staff participants emphasised multiple referral routes into Op COURAGE, including self-referrals and referrals via a professional. One of them identified the technical issues with form submission, which is being reviewed to make the submission easier.

> *“…it’s a bit of a convoluted thing where they need to sign it and then scan it and send it back. (we) have just been working on activating and amending the online referral form. So basically, that’ll mean that all people in future will be able to just fill it out on the website and not have to download anything”-MDT staff*

Some partners and collaborators mentioned that self-referral forms are straightforward. Nevertheless, other specified the difficulties veterans face in understanding the form, which would require a professional’s help. One partner agency participant spoke about making the form convenient enough for someone intoxicated to complete (a commonly reported issue among veterans – please see study from Rhead et al., 2022).

> *“you’ve got to design it so that someone who’s extremely poorly not feeling good, on their phone, not on a laptop - and their phone and their phone screen is cracked because they sat on their phone and they dropped it because they’re drunk. And they can look at it and go.”-Partner agency staff*

#### 4.2 Referrals require details

All collaborator and partner agency participants described the referral process at London Op COURAGE as straightforward. They highlighted the service staff’s accessibility in discussing a client before making the referral. One external organisation participant spoke about the swift triage and assessment.

> *“I think your speed of triage is the thing that really is much better…You obviously dedicated more resources getting triage done…nice and early is probably really where you’re acing this, because at least they’ve spoken to someone…if you’re dedicating extra resources to that…I think it’s a brilliant thing you’re doing.”-External organisation staff*

Partner agencies and collaborator participants suggested that the referrals received from the service need to be detailed, including information on any other aspect (such as housing) that the service is supporting them with. They also highlighted the referral to have information on the general practitioners’ advice and the veteran’s feasibility of using the internet.

> *“I think the referral process from Op COURAGE is patchy, to be honest… I think it not necessarily just a London issue, but across the country we often have challenges withsome of the referrals that are coming in as to whether they’re appropriate, whether certain conversations have been taken place with a veteran before being signposted to us.” - External organisation staff*

### 5. (In)visibility of the service and advertisement

Most partner agencies and external collaborator participants reported that the service may not be known to those serving or ex-military. They underscored the importance of advertising the service.

> *“some long termers that have been serving a lengthy sentence and will know nothing about Op COURAGE”-External organisation staff*

> *“I meet veterans that look blankly at me when I mention Operation COURAGE”-Partner agency staff*

There were opposing views among collaborator participants on the public awareness of the existence of the service. One of them said that anyone close to a veteran would know about the service, and another reported that a recent visit to a GP put forth the lack of awareness about the service. Most participants suggested that there is a need to advertise the service using posters in GP surgery waiting areas, improving veteran awareness among GPs, posting on social media and presenting at conferences and events.

> *“It’s websites, it’s Facebook, it’s getting out…with the posters and the banners. Turning up to different conferences or events…”-External organisation staff*

> *“Anyone that’s in the world of veterans and people’s well-being, I think they would have heard about Op COURAGE in some shape or form throughout the country.”-External organisation staff*

## Discussion

In this qualitative study, we aimed to explore the treatment and support offered by London Op COURAGE from the perspectives of professionals within and outside the service, supporting veterans and their families using the service. We gained insights from a diverse group of staff, which included multidisciplinary team members, partner agencies and external collaborators. We found that most professionals were impressed by the range and quality of resources available within the service. However, there were suggestions to improve the service’s communication, updates, and referral processes, as well as to increase its visibility and offer more veteran-specific training.

Veterans and their significant others are recognised as having distinct health needs [14]. Op COURAGE currently serves as a key veteran-sensitive point of access for veterans’ mental health and wellbeing needs in the UK. Our findings reveal that whilst the service is catering to the mental health of London-based veterans, they also work with charity partners and collaborators to offer holistic care for their service users. The wide array of requirements includes, but is not limited to, housing, employment, transition to civilian life and pension and compensation [7]. Our participants appreciated the service for having a holistic approach but were sometimes critical of not being updated regularly about service changes and their clients. The London Op COURAGE team could consider regular newsletters, meetings or social events to update their partners and collaborators as well as instil the feeling of inclusion among them.

To understand how multiple organisations collaborate in healthcare, there is a four-dimensional model [15]. The authors postulated that four elements are fundamental for the collaboration to be effective: (1) shared goals and vision, (2) a sense of belonging and mutual knowledge, (3) clear expectations and responsibilities, and finally (4) a leadership that encourages collaboration [15]. As per this model, internalisation (i.e. interdependencies and their management) and formalisation (i.e. communication and documentation) at London Op COURAGE could be strengthened further. There is also scope to employ specific link workers with particular external agencies.

Some of our participants raised the implications of non-veteran staff working in a service for veterans’ mental health and wellbeing. This is unlike the views of veteran service users and their family members, who, in previous research with the London Op COURAGE service, expressed that staff in the service are perceived as veteran-sensitive [16]. In this present study, an external collaborator suggested continuing professional development in the area of understanding the ranks and demands of veteran life. We propose ensuring that the staff are up to date with training and promoting veteran-specific knowledge and further involving veteran peer workers, deepening knowledge about military culture. Developing the service could also include highlighting the existing veteran staff to current and future clients as well as partner and external agencies. The service could also explore avenues to further involve veterans in the service in administration and operations, even if veteran mental health professionals are uncommon. Mandatory training using the existing online e-learning package, co-developed with veterans, for the service staff for veteran sensitive practice could be helpful.

### Strengths and Limitations

Our study had three key strengths. As a team of clinicians, academics and a veteran researcher, with consultation from service user advisors, we ensured that there were diverse and ground-level perspectives as well as methodological expertise throughout this study. We employed a qualitative approach to explore multiple stakeholder views on the treatment and support being offered at this service. The interviews and analysis were conducted by an independent team based at University College London (UCL), improving the likelihood of service providers being honest.

Our study nevertheless has some limitations. The service and partner agency staff may have been hesitant in participating and expressing their views, despite being interviewed by independent researchers. Online interviews have their merits but suffer from occasional technological issues, the lack of opportunity to observe non-verbal cues and potential initial rapport building. Finally, our use of purposive sampling could have limited the diversity of views. We would advise referring to staff perception cautiously. We have also reported service users’ and their significant others’ views of the London Op Courage [16].

## Conclusion

Our study highlighted staff perception of several strengths and some potential areas for improvement within the London Op COURAGE service. These interviews also elaborated on the multi-professional requirements of supporting a veteran at a specialist service such as the London Op COURAGE service. The partner and collaborator staff appreciated the convenience and accessibility of the service and their developing partnerships with the service staff. Improvements could be made in awareness about the service, continued development of the service staff and enhancing service user involvement. Nevertheless, addressing veterans’ multicomponent needs should take precedence in taking decisions about staff recruitment and service goals.

## Data Availability

Interview data has not been made publicly available in line with ethical requirements and the personal nature of the accounts shared by participants. If you need any information on the data, please contact the corresponding author, Prof Jo Billings.

## Acknowledgements

We would like to thank both lived experience advisors, Eric Crossfield and Anthony Muckell, who reviewed the interview questions.

---

**S1 Text** Topic Guide for interviews

